# Comparative performance of multiplex salivary and commercially available serologic assays to detect SARS-CoV-2 IgG and neutralization titers

**DOI:** 10.1101/2021.01.28.21250717

**Authors:** Christopher D. Heaney, Nora Pisanic, Pranay R. Randad, Kate Kruczynski, Tyrone Howard, Xianming Zhu, Kirsten Littlefield, Eshan U. Patel, Ruchee Shrestha, Oliver Laeyendecker, Shmuel Shoham, David Sullivan, Kelly Gebo, Daniel Hanley, Andrew D. Redd, Thomas C. Quinn, Arturo Casadevall, Jonathan M. Zenilman, Andrew Pekosz, Evan M. Bloch, Aaron A. R. Tobian

**Affiliations:** Department of Environmental Health and Engineering, Bloomberg School of Public Health, Johns Hopkins University, Baltimore, MD, USA; Department of Epidemiology, Bloomberg School of Public Health, Johns Hopkins University, Baltimore, MD, USA; Department of International Health, Bloomberg School of Public Health, Johns Hopkins University, Baltimore, MD, USA; Department of Pathology, Johns Hopkins University School of Medicine, Baltimore, MD, USA; Department of Molecular Microbiology and Immunology, Bloomberg School of Public Health, Johns Hopkins University, Baltimore, MD, USA; Department of Medicine, Johns Hopkins University School of Medicine, Baltimore, MD, USA; Department of Neurology, Johns Hopkins University School of Medicine, Baltimore, MD, USA; Division of Intramural Research, National Institute of Allergy and Infectious Diseases, National Institutes of Health, Baltimore MD

**Author notes:** **Corresponding author:** Christopher D. Heaney, PhD, MS, 615 North Wolfe Street, Room W7033B, Department of Environmental Health and Engineering, Bloomberg School of Public Health, Johns Hopkins University, Baltimore, MD, 21205, USA. Shared Senior Authors (Evan M. Bloch, Aaron A. R. Tobian). **Conflicts of interest:** The authors declare no potential conflicts of interest.

**Keywords:** Saliva, oral fluid, multiplex assay, SARS-CoV-2, COVID-19, serologic assays, neutralizing antibody, convalescent plasma

## Abstract

Oral fluid (hereafter saliva) offers a non-invasive sampling method for the detection of SARS-CoV-2 antibodies. However, data comparing performance of salivary tests against commercially-available serologic and neutralizing antibody (nAb) assays are lacking. This study compared the performance of a multiplex salivary SARS-CoV-2 IgG assay targeting antibodies to nucleocapsid (N), receptor binding domain (RBD) and spike (S) antigens to three commercially-available SARS-CoV-2 serology enzyme immunoassays (EIAs) (Ortho Vitros, Euroimmun, and BioRad) and nAb. Paired saliva and plasma samples were collected from 101 eligible COVID-19 convalescent plasma (CCP) donors >14 days since PCR+ confirmed diagnosis. Concordance was evaluated using positive (PPA) and negative (NPA) percent agreement, overall percent agreement (PA), and Cohen’s kappa coefficient. The range between salivary and plasma EIAs for SARS-CoV-2-specific N was PPA: 54.4-92.1% and NPA: 69.2-91.7%, for RBD was PPA: 89.9-100% and NPA: 50.0-84.6%, and for S was PPA: 50.6-96.6% and NPA: 50.0-100%. Compared to a plasma nAb assay, the multiplex salivary assay PPA ranged from 62.3% (N) and 98.6% (RBD) and NPA ranged from 18.8% (RBD) to 96.9% (S). Combinations of N, RBD, and S and a summary algorithmic index of all three (N/RBD/S) in saliva produced ranges of PPA: 87.6-98.9% and NPA: 50-91.7% with the three EIAs and ranges of PPA: 88.4-98.6% and NPA: 21.9-34.4% with the nAb assay. A multiplex salivary SARS-CoV-2 IgG assay demonstrated comparable performance to three commercially-available plasma EIAs and a nAb assay, and may be a viable alternative to assist in screening CCP donors and monitoring population-based seroprevalence and vaccine antibody response.

## INTRODUCTION

As coronavirus disease 2019 (COVID-19) emerged, there were limited diagnostic and treatment options; COVID-19 convalescent plasma (CCP) emerged early as one of the leading therapies. On August 23, 2020, the U.S. Food and Drug Administration (FDA) issued Emergency Use Authorization (EUA) approval for CCP; by December 2020 over 25,000 units of CCP were being transfused weekly in the United States (1, 4). According to the EUA, CCP units are required to be labeled as either high or low titer (11). This distinction is based on the signal to cut-off (S/C) ratio using a single assay i.e. the Ortho Vitros SARS-CoV-2 IgG assay. High and low titer is based on a signal to cutoff (S/CO) of ≥12 or <12 respectively (11).

Antibody titers can be determined by testing blood using commercially available enzyme immunoassays (EIAs) that typically measure antibody responses to a single antigen. Alternatively, microneutralization assays can be employed to determine a neutralizing antibody (nAb) titer. However, microneutralization requires both intensive biosecurity measures and substantial time, which are not amenable to high throughput donor screening.

Antibodies to SARS-CoV-2 have been evaluated in oral fluid (hereafter saliva), but little is known about how antibody titers in saliva correlate with those measured using plasma serologic assays for detection of SARS-CoV-2-specific IgG and nAb activity (12, 21, 25). If comparable performance were to be shown, saliva would offer several advantages over blood-based testing: collection is non-invasive and can be self-administered. These advantages would improve the scale and efficiency of CCP donor screening, population-based surveillance and assessment of vaccine responsiveness. This study sought to evaluate the performance of a multiplex salivary SARS-CoV-2 IgG assay relative to three commercially-available EIAs, and a nAb assay.

## METHODS

### Ethics statement

This study used stored samples and data from two parent studies that were approved by The Johns Hopkins University School of Medicine Institutional Review Board. All samples were de-identified prior to laboratory testing, and all participants provided informed consent. Both studies were conducted according to the ethical standards of the Helsinki Declaration of the World Medical Association.

### Study specimens

The stored plasma specimens that were used in this study had been collected from a convenience sample of potential CCP donors. The donors were recruited in the greater Baltimore, MD and Washington D.C. metropolitan areas from April to December 2020 (8, 15, 20). Saliva collection was undertaken in this cohort, starting in June 2020. Individuals were eligible for enrollment if they had a documented history of a positive molecular assay test result for SARS-CoV-2 infection (confirmed by medical chart review or the donor provided clinical documentation) and met standard self-reported eligibility criteria for blood donation. Only individuals who had both plasma and saliva collected on the same day were included in this study (n=108). The study used a complete case analysis approach, whereby 5 samples with missing values and 2 that did not pass QC were not used. Thus, 101 paired samples were included in the analysis. The study was cross-sectional and none of the subjects contributed more than one paired saliva / serum sample. All plasma samples were stored at −80°C until testing was performed.

Saliva was collected using the OraSure® Oral Antibody Collection Device (OraSure Technologies, Inc., Bethlehem, PA, USA). The saliva sample was processed according to manufacturer’s instructions, which involves adding the saliva contained in the Oral Antibody Collection Device foam paddle into 800 μL of OraSure® sample storage buffer immediately after the collection from participants. All samples were heated to 56°C for 1 hour to inactivate SARS-CoV-2 and stored at −80°C until analyzed. Archived pre-pandemic negative saliva samples were collected using the Oracol+ S14 collection device (Malvern Medical Developments, Ltd, Worcester, United Kingdom). These Oracol+ S14 samples were collected in multiple research studies prior to December 2019 and involved adult participants representing a diverse range of sociodemographic characteristics (18). Pre-pandemic negative saliva samples from prior to December 2019 were also heat-inactivated prior to testing with the multiplex assay.

### SARS-CoV-2 EIAs

Plasma specimens were analyzed using three commercially available EIAs: the Euroimmun Anti-SARS-CoV-2 ELISA (Mountain Lakes, NJ), the BioRad Platelia SARS-CoV-2 Total Ab (BioRad Laboratories, Hercules, CA), and the Ortho Vitros SARS-CoV-2 IgG EIA (Ortho-Clinical Diagnostics Inc, Rochester, NY). All EIAs were purchased from the manufacturer and conducted according to the manufacturers’ instructions, except for the Ortho Vitros EIA in which plasma—rather than serum as recommended—was used. The BioRad EIA measures IgG, IgM and IgA specific for SARS-CoV-2 nucleocapsid protein, whereas the Euroimmun and Ortho Vitros EIAs only measure anti-SARS-CoV-2 IgG specific to spike (S). The EIAs results are reported as follows: the Euroimmun EIA provides an arbitrary unit ratio (AU, which is the optical density [OD] of the sample divided by calibrator provided), the Ortho Vitros EIA provides a S/CO ratio, and the BioRad EIA provides an OD.

### Microneutralization Assay

Quantitation of nAb titers against 100 fifty percent tissue culture infectious doses (TCID50) was performed using a nAb assay (15). The nAb area under the curve (AUC) values were estimated using the exact number of wells protected from infection at every plasma dilution; samples that had no NT activity were assigned an arbitrary value of one-half of the lowest nAb AUC.

### Total salivary IgG ELISA

The total IgG concentration in each participant’s collected saliva (i.e., saliva added to OraSure sample storage buffer) was determined using Salimetrics Salivary Human Total IgG ELISA Kit (Salimetrics, LLC, Carlsbad, CA, USA) according to the manufacturer’s instructions with the following modification: the sample incubation and the detection antibody incubation times were reduced to 1 hour instead of 2 hours. This modification was approved by the manufacturer.

### Multiplex SARS-CoV-2 IgG test

Saliva samples were tested using a multiplex SARS-CoV-2 IgG immunoassay as previously described, based on Luminex technology (21). The multiplex assay included magnetic bead sets (MagPlex microspheres) coupled covalently with antigen (5 μg antigen per 1 million beads) (21). The assay included SARS-CoV-2 nucleocapsid (N), receptor binding domain (RBD), and spike (S) antigens (**Supplemental Table 1**). Briefly, saliva was centrifuged for 5 minutes at 20,000 g. Because all CCP saliva samples were collected using the OraSure® Oral Antibody Collection Device (OraSure Technologies, Inc., Bethlehem, PA, USA), which includes 800 μL of OraSure® sample storage buffer, the saliva contained in the collection device (foam paddle) is diluted (estimated 4-fold dilution compared to our prior testing of Oracol-collected saliva samples). Thus, instead of 10 μL as described previously (21), 40 μL of this combined saliva / OraSure sample buffer was added to each well along with 10 μL PBST/1% BSA (assay buffer) containing 1,000 beads per bead set for a final volume of 50 μL. Each assay plate contained 1-2 blank wells with OraSure sample buffer instead of samples that were used for background subtraction. A positive control was created by spiking a during-pandemic saliva sample that was highly positive for SARS-CoV-2-specific IgG into a pre-pandemic negative control saliva sample. The same pre-pandemic saliva was used as a negative control. Because the pre-pandemic saliva samples were collected using the Oracol+ S14 device, which does not contain any sample storage buffer, 10 μL of undiluted saliva was added to each well along with 40 μL PBST/1% BSA (assay buffer) containing 1,000 beads per bead set for a final volume of 50 μL. Phycoerythrin-labeled anti-human IgG diluted 1:100 in assay buffer was used to detect IgG binding to antigens. The plates were read on a Luminex MAGPIX instrument.

**Table 1.** **Concordance between salivary SARS-Cov-2 antibody assays and serology antibody tests in COVID-19 convalescent individuals**.

| Salivary assay<sup>a</sup> | Ortho Vitros (88+, 13-)<sup>b</sup> |  |  |  | Euroimmun (90+, 11-)<sup>b</sup> |  |  |  | BioRad (89+, 12-)<sup>b</sup> |  |  |  | nAb (69+, 32-)<sup>c</sup> |  |  |  |
| --- | --- | --- | --- | --- | --- | --- | --- | --- | --- | --- | --- | --- | --- | --- | --- | --- |
|  | PPA | NPA | PA | Kappa | PPA | NPA | PA | Kappa | PPA | NPA | PA | Kappa | PPA | NPA | PA | Kappa |
| GenScript N<sup>d</sup> | 55.7% | 84.6% | 59.4% | 0.182 | 54.4% | 81.8% | 57.4% | 0.142 | 56.2% | 91.7% | 60.4% | 0.202 | 62.3% | 75.0% | 66.3% | 0.324 |
| NAC N<sup>d</sup> | 89.8% | 69.2% | 87.1% | 0.507 | 88.9% | 72.7% | 87.1% | 0.482 | 92.1% | 91.7% | 92.1% | 0.689 | 89.9% | 34.4% | 72.3% | 0.274 |
| Sino Bio RBD<sup>d</sup> | 100.0% | 53.8% | 94.1% | 0.670 | 98.9% | 54.5% | 94.1% | 0.636 | 98.9% | 50.0% | 93.1% | 0.596 | 98.6% | 18.8% | 73.3% | 0.219 |
| Mt. Sinai RBD<sup>d</sup> | 93.2% | 84.6% | 92.1% | 0.688 | 91.1% | 81.8% | 90.1% | 0.588 | 89.9% | 66.7% | 87.1% | 0.479 | 91.3% | 34.4% | 73.3% | 0.294 |
| GenScript RBD<sup>d</sup> | 97.7% | 76.9% | 95.0% | 0.772 | 95.6% | 72.7% | 93.1% | 0.657 | 95.5% | 66.7% | 92.1% | 0.622 | 94.2% | 25.0% | 72.3% | 0.231 |
| Sino Bio ECD<sup>d</sup> | 96.6% | 53.8% | 91.1% | 0.559 | 95.6% | 54.5% | 91.1% | 0.522 | 95.5% | 50.0% | 90.1% | 0.490 | 97.1% | 25.0% | 74.3% | 0.271 |
| Mt. Sinai S<sup>d</sup> | 52.3% | 100.0% | 58.4% | 0.220 | 51.1% | 100.0% | 56.4% | 0.185 | 50.6% | 91.7% | 55.4% | 0.166 | 65.2% | 96.9% | 75.2% | 0.521 |
| Sum N S/CO<sup>e</sup> | 89.8% | 69.2% | 87.1% | 0.507 | 88.9% | 72.7% | 87.1% | 0.482 | 92.1% | 91.7% | 92.1% | 0.689 | 89.9% | 34.4% | 72.3% | 0.274 |
| Sum RBD S/CO<sup>f</sup> | 98.9% | 76.9% | 96.0% | 0.811 | 96.7% | 72.7% | 94.1% | 0.694 | 96.6% | 66.7% | 93.1% | 0.657 | 95.7% | 25.0% | 73.3% | 0.251 |
| Sum S S/CO<sup>g</sup> | 89.8% | 61.5% | 86.1% | 0.454 | 87.8% | 54.5% | 84.2% | 0.341 | 87.6% | 50.0% | 83.2% | 0.319 | 88.4% | 28.1% | 69.3% | 0.189 |
| Sum N/RBD/S S/CO<sup>h</sup> | 98.9% | 53.8% | 93.1% | 0.630 | 97.8% | 54.5% | 93.1% | 0.594 | 98.9% | 58.3% | 94.1% | 0.668 | 98.6% | 21.9% | 74.3% | 0.256 |

Note: S/CO: signal to cut-off ratio; PPA: positive percent agreement; NPA: negative percent agreement; PA: percent agreement; Kappa: Cohen's kappa coefficient; nAb: Neutralizing antibody; NAC:
Native Antigen Company; Sino Bio: Sino Biological
<sup>a</sup>Insufficient total IgG, instrument error, ID discrepancy or other exclusion reasons for Final Result were set to missing.
<sup>b</sup>Tests were considered positive result (+) or negative result (-) per manufacturer's cutoff. Indeterminate results for blood antibody tests were considered negative.
<sup>c</sup>nAb area under curve (AUC) <20 was considered as negative result (-), ≥20 as positive result (+).
<sup>d</sup>Each salivary assay result was considered positive, if median fluorescence intensity (MFI) above cut-off, and considered negative if MFI below cut-off.
<sup>e</sup>Sum N S/CO was considered positive if the sum of signal to cutoff ratios of 2 N antigens above threshold (mean plus 3 SD of pre-COVID samples that have >3.75 ug/mL total IgG).
<sup>f</sup>Sum RBD S/CO was considered positive if the sum of signal to cutoff ratios of 5 RBD antigens above threshold (mean plus 3 SD of pre-COVID samples that have >3.75 ug/mL total IgG).
<sup>g</sup>Sum S S/CO was considered positive if the sum of signal to cutoff ratios of 2 antigens above threshold (mean plus 3 SD of pre-COVID samples that have >3.75 ug/mL total IgG).
<sup>h</sup>Sum N/S/RBD S/CO was considered positive if the sum of 2 N, 3 RBD and 2 S antigen signal to cutoff values > 6.

### Statistical analysis

For salivary SARS-CoV-2 multiplex assay results, the blank-subtracted (“net”) median fluorescence intensity (MFI) was used for statistical analyses. Cutoffs to discriminate IgG positive from IgG negative samples for each individual antigen and for combinations of multiple antigens (algorithms) had previously been calculated using the average net MFI plus three standard deviations of pre-COVID-19 era negative control specimens (n=265). The sensitivity and specificity for individual and for combinations of SARS-CoV-2 antigens had been calculated using saliva samples collected >14 days after COVID-19 symptoms onset and pre-COVID-19 era saliva samples as described elsewhere (21)]. Highest accuracy (98.6% sensitivity [143/145] and 99.2% specificity [263/265] was achieved using a summary index of SARS-CoV-2-specific IgG S/CO to seven N, RBD and S antigens [unpublished data] and by applying a minimum sample quality control (QC) threshold based on total salivary IgG concentration (μg/mL). A total salivary IgG QC threshold was applied to samples that were negative for the summary index of SARS-CoV-2-specific IgG S/CO to seven N, RBD and S antigens. As with pre-pandemic Oracol-collected saliva samples, any OraSure-collected saliva sample containing less than 0.15 μg total IgG per 50-μL assay reaction was considered to not pass sample QC and was excluded from the analysis if the summary index of SARS-CoV-2-specific IgG S/CO to seven N, RBD and S antigens did not cross the cutoff of 6. This quality control measure excludes samples that could potentially be classified as false negatives as a result of improper saliva sample collection or insufficient total salivary IgG concentration.

The concordance of the multiplex salivary SARS-Cov-2 IgG assay with 3 blood-based EIAs (using manufacturer’s cutoffs) and nAb titers (using AUC of 20 as cutoff) was examined using positive percent agreement (PPA), negative percent agreement (NPA), percent agreement (PA) and Cohen’s kappa coefficient. Indeterminate and borderline results of Euroimmun and BioRad were considered to be negative. Spearman’s rank correlation coefficients (ρ) were used to examine the correlation of the multiplex salivary EIA’s signal to cut off (S/CO) and blood-based test values (Ortho Vitros S/CO, Euroimmun AU, BioRad OD and nAb AUC); 95% confidence intervals (CI) were estimated by 1,000 bootstrap iterations. Concordance of each antigen-specific component of the salivary assay and the final algorithmic result were examined to evaluate the driving component of the final result. The concordance and correlation between each blood-based test were also examined. To calculate the area under the receiver operating characteristic (AUROC), receiver operating characteristic (ROC) analysis for the multiplex salivary summary index S/CO ratio for N/RBD/S at various thresholds to detect SARS-CoV-2 high antibody titers were performed. Statistical analyses were conducted in R 4.0.3 (R Core Team, Vienna, Austria).

## RESULTS

### Specimen characteristics

Demographic information for the subjects that contributed specimens for this analysis is shown in **Supplemental Table 2** (n=101). The median age was 44 years (interquartile range [IQR]=34-56), 42.6% were male, and the majority of the participants (72.3%) were non-Hispanic White. Only 14.9% were hospitalized due to COVID-19. There was a median of 50 days (IQR=40-70 days) between diagnostic PCR+ assay and sample collection for this study.

### Comparative performance of saliva to detect antibodies to SARS-CoV-2

In comparison to the three commercial serological EIAs, performance was generally best for the RBD antigens within the multiplex salivary SARS-CoV-2 IgG assay and the Ortho Vitros EIA (**Table 1**). The highest percent agreement was between the Ortho Vitros EIA and the multiplex salivary SARS-CoV-2 IgG assay’s GenScript RBD-specific IgG (PPA=97.7%; NPA=76.9%; PA=95.0%) and sum of all three RBD-specific IgG S/CO values (PPA=98.9%, NPA=76.9%; PA=96.0%) (**Table 1**). The sum of all N, S, and RBD antigen S/CO values (summary index) also demonstrated good percent agreement with the Ortho Vitros EIA (PPA=98.9%; NPA=53.8; PA=93.1%) (**Table 1**). Good percent agreement was also observed between the multiplex salivary SARS-CoV-2 IgG assay’s RBD antigens and the Euroimmun (PPA range: 91.1 %-98.9%; NPA range: 54.5%-81.8%; PA range: 90.1%-94.1%) and BioRad (PPA range: 89.9%-98.9%; NPA range: 50.0%-66.7%; PA range: 87.1%-93.1%) serological EIAs (**Table 1**).

In comparison to a nAb assay (considering an area under the curve [AUC] <20 as a negative result and ≥20 as a positive result), the best performance was observed for the multiplex salivary SARS-CoV-2 IgG assay’s Mt. Sinai whole spike-specific IgG result (PPA=65.2%; NPA=96.9%; PA=75.2%). The next best comparative performance with nAb was followed closely by the multiplex salivary SARS-CoV-2 IgG assay’s Sino Bio ECD antigen (PPA=97.1%; NPA=25.0%; PA=74.3%) and the NAC N and all 3 RBD antigens (PA range: 72.3%-73.3%) (**Table 1**). The concordance between each component of the multiplex SARS-CoV-2 antibody assay with the final algorithmic result in the multiplex salivary assay is presented in **Supplemental Table 3**.

The concordance between SARS-CoV-2 plasma antibody tests is shown in **Supplemental Table 4**. The comparative performance of the Ortho Vitros EIA (PPA=75.0%; NPA=76.9%; PA=75.2%), Euroimmun EIA (PPA=98.6%; NPA=31.3%; PA=77.2%) and BioRad EIA (PPA=95.7%; NPA=28.1%; PA=74.3%) with nAb was similar to several of the antigens in the multiplex salivary SARS-CoV-2 IgG assay, particularly Mt. Sinai whole spike, all 3 RBDs and NAC N. The comparative performance of the multiplex salivary SARS-CoV-2 assay with Ortho Vitros EIA was slightly lower than that of the Euroimmun EIA with the OrthoVitros EIA (PPA=100.0%; NPA=84.6%; PA=98.0%) but higher than that of the BioRad EIA with OrthoVitros EIA (PPA=95.5%; NPA=61.5%; PA=91.1%)

The multiplex SARS-CoV-2 IgG assay’s three RBD antigens and Mt. Sinai whole S antigen demonstrated the highest correlations with the three commercially available serological EIAs and nAb (Spearman rank correlation coefficient [ρ] range for Ortho Vitros=0.81-0.86, Euroimmun=0.79-0.83, BioRad=0.39-0.44, and nAb=0.75-0.77) (**Supplemental Table 5**). The integrated sum of all three RBD S/CO values was most strongly correlated with the Ortho Vitros EIA (ρ=0.86; 95% confidence interval [CI]=0.76, 0.91; *p*<0.001), followed by Euroimmun (ρ=0.83; 95% CI=0.74, 0.89; *p*<0.001) and nAb (ρ=0.77; 95% CI=0.66, 0.85; *p*<0.001) (**Figure 1**). Correlations between SARS-CoV-2 plasma antibody tests are shown in **Supplemental Table 6**. Correlations with nAb AUC were better for the Ortho Vitros EIA S/CO (ρ=0.83; 95% CI=0.74, 0.89; *p*<0.001) and Euroimmun EIA AU (ρ=0.80; 95% CI=0.72, 0.86; *p*<0.001) than those for the multiplex salivary assay and nAb AUC.

**Figure 1.**
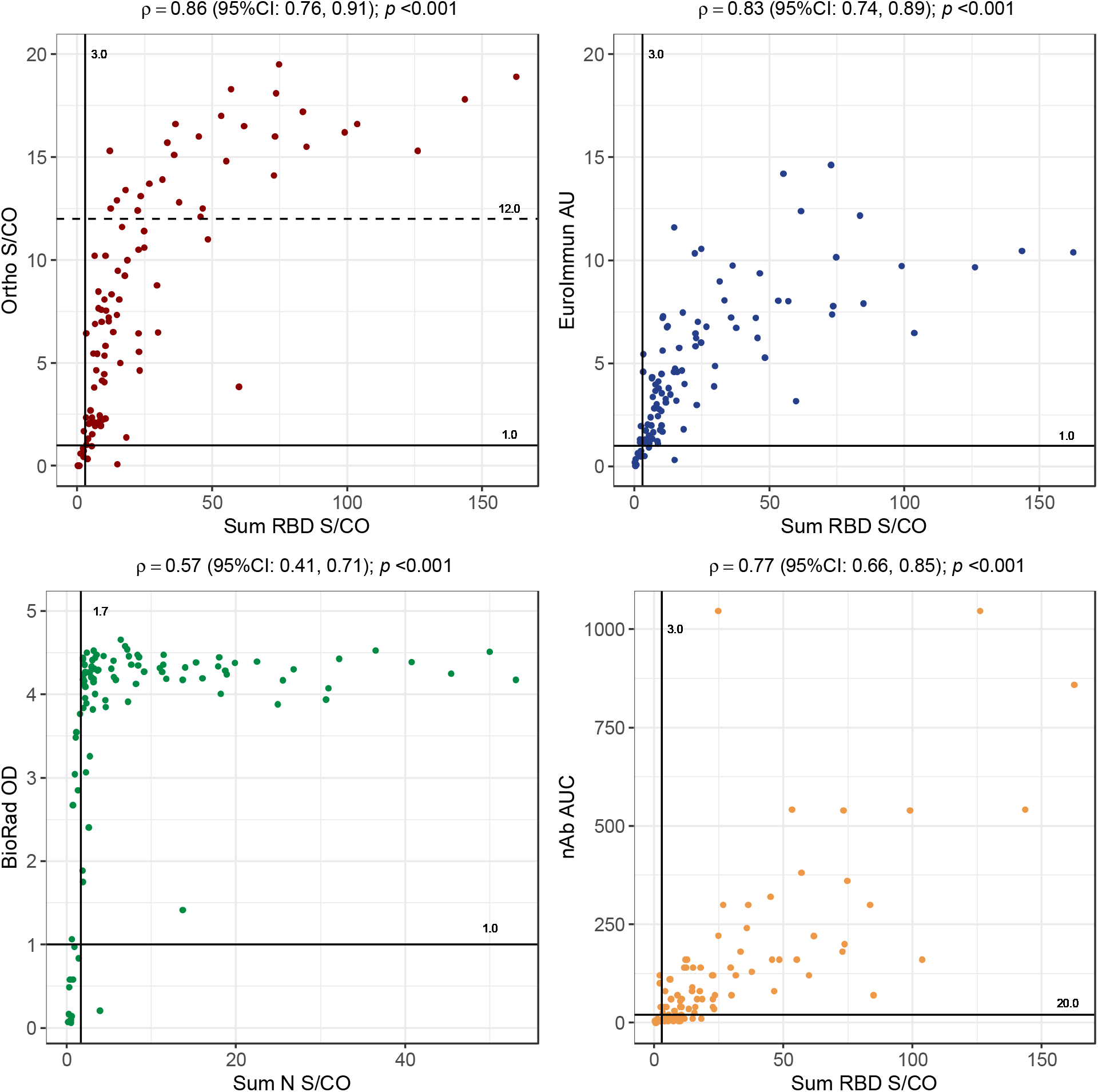
Correlations between saliva SARS-Cov-2 antibody assay and serology antibody tests in COVID-19 convalescent individuals. Note: S/CO = signal to cut off ratio. AU = arbitrary units. OD = optical density. nAb = neutralizing antibody. AUC: area under curve. Only the saliva assays having the highest Spearman’s correlation coefficients (ρ) with each blood assay were plot. ρ were calculated with 95% confidence intervals (CI) estimated over 1000 bootstrap iterations. The straight vertical black line indicates the cut-off for SARS-CoV-2 positivity of saliva assays. The straight horizontal black line represents the cut-off for SARS-CoV-2 positivity of serology assays. The dashed horizontal line indicates the cut-off for SARS-Cov-2 high antibody titer for Ortho Vitros test.

When compared to a S/CO value of 12 or greater using the Ortho Vitros EIA (i.e., the requirement for high-titer designation under the EUA), the receiver operating characteristic (ROC) area under the curve (AUC) for the multiplex salivary assay’s sum of S/CO ratios for N/RBD/S antigens was 0.92 (**Figure 2**).

**Figure 2.**
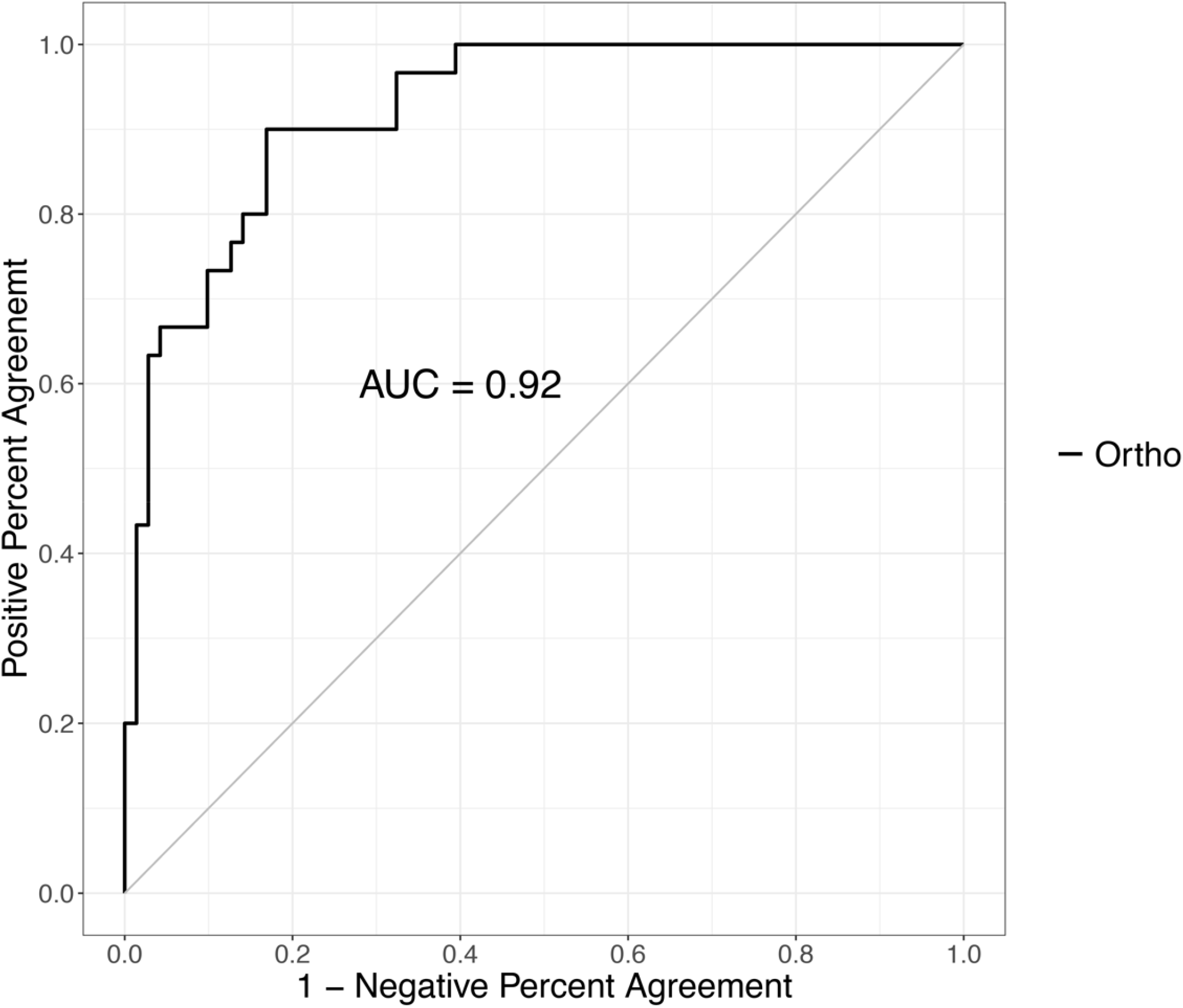
Receiver operating characteristic (ROC) curve for multiplex salivary SARS-CoV-2 IgG assay’s sum of the signal to cut-off ratio for N/RBD/S at various thresholds to detect SARS-CoV-2 high antibody titers. Note: a signal to cut-off value of 12 or greater of Ortho Vitros EIA were considered high antibody titer. AUC: area under curve.

## DISCUSSION

In this study of eligible CCP donors, a multiplex salivary SARS-CoV-2 IgG assay’s performance was comparable to three commercially-available SARS-CoV-2 serological tests that are commonly used to qualify high-titer CCP and as surrogates of nAb activity. Importantly, as a surrogate of nAb activity, the multiplex salivary SARS-CoV-2 IgG assay appeared to demonstrate equivalent or slightly better percent agreement than the three commercially-available serological EIAs. The high comparative performance of the salivary multiplex SARS-CoV-2 IgG assay is evident in the higher percent agreement for S components of the multiplex and the Ortho Vitros and Euroimmun EIAs (which both measure S-specific IgG responses) and N components of the multiplex and the BioRad EIA (which measures N-specific IgG responses).

Saliva has become an important specimen type for the diagnosis of both active and previous infection with SARS-CoV-2. For one, saliva collection is minimally-invasive when compared to other commonly used sampling approaches such as phlebotomy and nasopharyngeal-, mid-turbinate-, or anterior nares swabs. Saliva has been shown to be a robust alternative to nasopharyngeal swabs for SARS-CoV-2 RNA testing, which lends itself to population-level diagnosis and/or surveillance (6, 19, 28, 29). Specifically, collection can be self-administered without technical expertise or oversight. A recent study suggested that salivary SARS-CoV-2 viral load can serve as a dynamic correlate of COVID-19 severity and mortality(24). The utility of SARS-CoV-2 RNA detection in saliva has emerged in parallel with proofs-of-principle to detect SARS-CoV-2-specific antibody responses in saliva, which could serve as a surrogate of SARS-CoV-2 serological tests (10, 12, 17, 21). SARS-CoV-2-specific IgG in saliva reflects the blood-derived transudate in the oral cavity; by contrast, SARS-CoV-2-specific IgA in saliva may represent a localized mucosal IgA response to infection (2, 22, 27).

Because salivary matrix effects are complex and IgG antibodies are present at lower concentrations than in blood, maintaining the diagnostic accuracy – particularly high sensitivity or PPA – of a salivary “serological” assay relative to blood-based serological assays has proven challenging. Our multiplex assay approach produces robust signals for combinations of SARS-CoV-2 N, RBD, and S antigens within a single saliva sample. This facilitates optimization of algorithms that can produce both high sensitivity and specificity(21). Furthermore, the semi-quantitative nature of the S/CO values generated by the salivary SARS-CoV-2 multiplex IgG assay could offer insight into factors driving the duration and magnitude of SARS-CoV-2 IgG seropositivity in high risk and general populations (5, 7, 9, 23).

Saliva can be used to investigate vaccine immunogenicity (i.e., responsiveness) and population seroprevalence. It can also be beneficial for screening of donors for collection of CCP. In the US, CCP has emerged as one of the major treatments for those hospitalized with COVID-19, whereby efficacy has been demonstrated, particularly when units are high-titer and are administered early in the disease course (13, 16, 26). Despite rapid scale up of CCP collection in 2020, enabling procurement of 25,000-30,000 units of CCP per week, the national inventory of CCP is declining, owing to ongoing clinical demand. Clinical trials are currently underway to evaluate the efficacy of CCP as post-exposure prophylaxis and early treatment. If those trials show positive results, the demand for CCP could increase significantly. In addition, the FDA requires the labelling units of CCP as high- or low-titer under the EUA; this further detracts from CCP inventories given that only a subset of donors will satisfy criteria for high-titer. Given the ease of collection, saliva can be used to enhance screening, thereby favorably impacting CCP collections.

This study has several limitations. First, the sample size was relatively small and the cross-sectional design precluded evaluation of antibody dynamics over time, particularly given the relatively short period during which sample collection was undertaken (i.e. relative to symptom resolution). Nonetheless, this is consistent with most CCP collections and study populations, which provided insight into SARS-CoV-2 immunopathogenesis and screening options(3, 14). Second, the study population was primarily focused around Baltimore, MD and Washington DC, thus potentially limiting generalizability of the findings. Third, the Ortho Vitros EIA was validated for serum rather than plasma; while we do not believe that this impacted the results, it is a departure from the manufacturer’s instructions. Finally, a different saliva collection device (OraSure, Bethlehem, PA, USA) was used to establish the negative threshold for the multiplex SARS-CoV-2 IgG assay (Oracol+ S14, Malvern Medical Developments, Worcester, UK). A larger study is recommended to test reproducibility of the findings following the manufacturer instructions for the assay kits and using the same saliva collection devices. Nonetheless, the study employed a robust multiplex saliva assay approach to detect antibodies against SARS-CoV-2, demonstrating favorable performance both against several commercial EIAs, including the Ortho Vitros, as well as formal viral neutralization.

When applied to saliva, the highly-adaptable multiplex approach enables detection of a diverse range of SARS-CoV-2 antigen-specific IgG responses using a minimally-invasive biospecimen that can be self-collected at home. If the findings are replicated in larger studies, this could support a large scale-up of potential CCP donor screening as well as of general population SARS-CoV-2 serosurveillance. This saliva-based approach could improve population-scale understanding of the extent of SARS-CoV-2 transmission, identify areas with gaps in immunity, and also support monitoring of the magnitude and duration of natural infection. These data may be used to guide vaccination decision-making.

## Supporting information

Supplemental Tables 1 through 6

## Data Availability

A de-identified dataset may be made available upon request.

## ACKNOWLEDGEMENTS

We acknowledge all of the participants who contributed specimens to this study and the study staff without whom this study would not have been possible. We thank Florian Krammer, Icahn School of Medicine at Mt. Sinai for providing S and S-RBD protein. We also thank the National Institute of Infectious Diseases, Japan, for providing VeroE6TMPRSS2 cells and acknowledge the Centers for Disease Control and Prevention, BEI Resources, NIAID, NIH for SARS-Related Coronavirus 2, Isolate USA-WA1/2020, NR-5228.

## REFERENCES

1. ABC 2020, posting date. Newsletter #44: COVID-19 Convalescent Plasma Updates. [Online.]

2. Aita, A., D. Basso, A. M. Cattelan, P. Fioretto, F. Navaglia, F. Barbaro, A. Stoppa, E. Coccorullo, A. Farella, A. Socal, R. Vettor, and M. Plebani. 2020. SARS-CoV-2 identification and IgA antibodies in saliva: One sample two tests approach for diagnosis. Clin Chim Acta 510:717–722.

3. Benner, S. E., E. U. Patel, O. Laeyendecker, A. Pekosz, K. Littlefield, Y. Eby, R. E. Fernandez, J. Miller, C. S. Kirby, M. Keruly, E. Klock, O. R. Baker, H. A. Schmidt, R. Shrestha, I. Burgess, T. S. Bonny, W. Clarke, P. Caturegli, D. Sullivan, S. Shoham, T. C. Quinn, E. M. Bloch, A. Casadevall, A. A. R. Tobian, and A. D. Redd. 2020. SARS-CoV-2 Antibody Avidity Responses in COVID-19 Patients and Convalescent Plasma Donors. J Infect Dis 222:1974–1984.

4. Bloch, E. M., S. Shoham, A. Casadevall, B. S. Sachais, B. Shaz, J. L. Winters, C. van Buskirk, B. J. Grossman, M. Joyner, J. P. Henderson, A. Pekosz, B. Lau, A. Wesolowski, L. Katz, H. Shan, P. G. Auwaerter, D. Thomas, D. J. Sullivan, N. Paneth, E. Gehrie, S. Spitalnik, E. A. Hod, L. Pollack, W. T. Nicholson, L. A. Pirofski, J. A. Bailey, and A. A. Tobian. 2020. Deployment of convalescent plasma for the prevention and treatment of COVID-19. J Clin Invest 130:2757–2765.

5. Bruni, M., V. Cecatiello, A. Diaz-Basabe, G. Lattanzi, E. Mileti, S. Monzani, L. Pirovano, F. Rizzelli, C. Visintin, G. Bonizzi, M. Giani, M. Lavitrano, S. Faravelli, F. Forneris, F. Caprioli, P. G. Pelicci, G. Natoli, S. Pasqualato, M. Mapelli, and F. Facciotti. 2020. Persistence of Anti-SARS-CoV-2 Antibodies in Non-Hospitalized COVID-19 Convalescent Health Care Workers. J Clin Med 9.

6. Butler-Laporte, B., A. Lawandi, I. Schiller, M. C. Yao, N. Dendukuri, E. G. McDonald, and T. C. Lee. 2021. Comparison of Saliva and Nasopharyngeal Swab Nucleic Acid Amplification Testing for Detection of SARS-CoV-2: A Systematic Review and Meta-analysis. JAMA Intern Med.

7. Chen, Y., A. Zuiani, S. Fischinger, J. Mullur, C. Atyeo, M. Travers, F. J. N. Lelis, K. M. Pullen, H. Martin, P. Tong, A. Gautam, S. Habibi, J. Bensko, D. Gakpo, J. Feldman, B. M. Hauser, T. M. Caradonna, Y. Cai, J. S. Burke, J. Lin, J. A. Lederer, E. C. Lam, C. L. Lavine, M. S. Seaman, B. Chen, A. G. Schmidt, A. B. Balazs, D. A. Lauffenburger, G. Alter, and D. R. Wesemann. 2020. Quick COVID-19 Healers Sustain Anti-SARS-CoV-2 Antibody Production. Cell 183:1496–1507 e16.

8. Conklin, S. E., K. Martin, Y. C. Manabe, H. A. Schmidt, J. Miller, M. Keruly, E. Klock, C. S. Kirby, O. R. Baker, R. E. Fernandez, Y. J. Eby, J. Hardick, K. Shaw-Saliba, R. E. Rothman, P. P. Caturegli, A. D. Redd, A. A. R. Tobian, E. M. Bloch, H. B. Larman, T. C. Quinn, W. Clarke, and O. Laeyendecker. 2020. Evaluation of Serological SARS-CoV-2 Lateral Flow Assays for Rapid Point of Care Testing. J Clin Microbiol.

9. Dan, J. M., J. Mateus, Y. Kato, K. M. Hastie, E. D. Yu, C. E. Faliti, A. Grifoni, S. I. Ramirez, S. Haupt, A. Frazier, C. Nakao, V. Rayaprolu, S. A. Rawlings, B. Peters, F. Krammer, V. Simon, E. O. Saphire, D. M. Smith, D. Weiskopf, A. Sette, and S. Crotty. 2021. Immunological memory to SARS-CoV-2 assessed for up to 8 months after infection. Science.

10. Faustini, S. E., S. E. Jossi, M. Perez-Toledo, A. Shields, J. D. Allen, Y. Watanabe, M. L. Newby, A. Cook, C. R. Willcox, M. Salim, M. Goodall, J. L. Heaney, E. Marcial-Juarez, G. L. Morley, B. Torlinska, D. C. Wraith, T. Veenith, S. Harding, S. Jolles, P. J. Mark, T. Plant, A. Huissoon, M. K. O’Shea, B. E. Willcox, M. T. Drayson, M. Crispin, A. F. Cunningham, and A. G. Richter. 2020. Detection of antibodies to the SARS-CoV-2 spike glycoprotein in both serum and saliva enhances detection of infection. medRxiv.

11. FDA. 2020. Emergency Use Authorization (EUA) 26382: EUA Request (original request 8/12/20; amended request 8/23/20). In CBER (ed.).

12. Isho, B., K. T. Abe, M. Zuo, A. J. Jamal, B. Rathod, J. H. Wang, Z. Li, G. Chao, O. L. Rojas, Y. M. Bang, A. Pu, N. Christie-Holmes, C. Gervais, D. Ceccarelli, P. Samavarchi-Tehrani, F. Guvenc, P. Budylowski, A. Li, A. Paterson, F. Y. Yue, L. M. Marin, L. Caldwell, J. L. Wrana, K. Colwill, F. Sicheri, S. Mubareka, S. D. Gray-Owen, S. J. Drews, W. L. Siqueira, M. Barrios-Rodiles, M. Ostrowski, J. M. Rini, Y. Durocher, A. J. McGeer, J. L. Gommerman, and A. C. Gingras. 2020. Persistence of serum and saliva antibody responses to SARS-CoV-2 spike antigens in COVID-19 patients. Sci Immunol 5.

13. Joyner, M. J., J. W. Senefeld, S. A. Klassen, J. R. Mills, P. W. Johnson, E. S. Theel, C. C. Wiggins, K. A. Bruno, A. M. Klompas, E. R. Lesser, K. L. Kunze, M. A. Sexton, J. C. Diaz Soto, S. E. Baker, J. R. A. Shepherd, N. van Helmond, C. M. van Buskirk, J. L. Winters, J. R. Stubbs, R. F. Rea, D. O. Hodge, V. Herasevich, E. R. Whelan, A. J. Clayburn, K. F. Larson, J. G. Ripoll, K. J. Andersen, M. R. Buras, M.N. P. Vogt, J. J. Dennis, R. J. Regimbal, P. R. Bauer, J. E. Blair, N. S. Paneth, D. Fairweather, R. S. Wright, R. E. Carter, and A. Casadevall. 2020. Effect of Convalescent Plasma on Mortality among Hospitalized Patients with COVID-19: Initial Three-Month Experience. medRxiv.

14. Kared, H., A. D. Redd, E. M. Bloch, T. S. Bonny, H. R. Sumatoh, F. Kairi, D. Carbajo, B. Abel, E. W. Newell, M. Bettinotti, S. E. Benner, E. U. Patel, K. Littlefield, O. Laeyendecker, S. Shoham, D. Sullivan, A. Casadevall, A. Pekosz, A. Nardin, M. Fehlings, A. A. Tobian, and T. C. Quinn. 2021. SARS-CoV-2-specific CD8+ T cell responses in convalescent COVID-19 individuals. J Clin Invest.

15. Klein, S. L., A. Pekosz, H. S. Park, R. L. Ursin, J. R. Shapiro, S. E. Benner, K. Littlefield, S. Kumar, H. M. Naik, M. J. Betenbaugh, R. Shrestha, A. A. Wu, R. M. Hughes, I. Burgess, P. Caturegli, O. Laeyendecker, T. C. Quinn, D. Sullivan, S. Shoham, A. D. Redd, E. M. Bloch, A. Casadevall, and A. A. Tobian. 2020. Sex, age, and hospitalization drive antibody responses in a COVID-19 convalescent plasma donor population. J Clin Invest 130:6141–6150.

16. Libster, R., G. Perez Marc, D. Wappner, S. Coviello, A. Bianchi, V. Braem, I. Esteban, M. T. Caballero, C. Wood, M. Berrueta, A. Rondan, G. Lescano, P. Cruz, Y. Ritou, V. Fernandez Vina, D. Alvarez Paggi, S. Esperante, A. Ferreti, G. Ofman, A. Ciganda, R. Rodriguez, J. Lantos, R. Valentini, N. Itcovici, A. Hintze, M. L. Oyarvide, C. Etchegaray, A. Neira, I. Name, J. Alfonso, R. Lopez Castelo, G. Caruso, S. Rapelius, F. Alvez, F. Etchenique, F. Dimase, D. Alvarez, S. S. Aranda, C. Sanchez Yanotti, J. De Luca, S. Jares Baglivo, S. Laudanno, F. Nowogrodzki, R. Larrea, M. Silveyra, G. Leberzstein, A. Debonis, J. Molinos, M. Gonzalez, E. Perez, N. Kreplak, S. Pastor Arguello, L. Gibbons, F. Althabe, E. Bergel, and F. P. Polack. 2021. Early High-Titer Plasma Therapy to Prevent Severe Covid-19 in Older Adults. N Engl J Med.

17. MacMullan, M. A., A. Ibrayeva, K. Trettner, L. Deming, S. Das, F. Tran, J. R. Moreno, J. G. Casian, P. Chellamuthu, J. Kraft, K. Kozak, F. E. Turner, V. I. Slepnev, and L. M. Le Page. 2020. ELISA detection of SARS-CoV-2 antibodies in saliva. Sci Rep 10:20818.

18. Nadimpalli, M. L., J. R. Stewart, E. Pierce, N. Pisanic, D. C. Love, D. Hall, J. Larsen, K. C. Carroll, T. Tekle, T. M. Perl, and C. D. Heaney. 2018. Face Mask Use and Persistence of Livestock-associated Staphylococcus aureus Nasal Carriage among Industrial Hog Operation Workers and Household Contacts, USA. Environ Health Perspect 126:127005.

19. Ott, I. M., M. S. Strine, A. E. Watkins, M. Boot, C. C. Kalinich, C. A. Harden, C. B. F. Vogels, A. Casanovas-Massana, A. J. Moore, M. C. Muenker, M. Nakahata, M. Tokuyama, A. Nelson, J. Fournier, S. Bermejo, M. Campbell, R. Datta, C. S. Dela Cruz, S. F. Farhadian, A. I. Ko, A. Iwasaki, N. D. Grubaugh, C. B. Wilen, and A. L. Wyllie. 2020. Simply saliva: stability of SARS-CoV-2 detection negates the need for expensive collection devices. medRxiv.

20. Patel, E. U., E. M. Bloch, W. Clarke, Y. H. Hsieh, D. Boon, Y. Eby, R. E. Fernandez, O. R. Baker, M. Keruly, C. S. Kirby, E. Klock, K. Littlefield, J. Miller, H. A. Schmidt, P. Sullivan, E. Piwowar-Manning, R. Shrestha, A. D. Redd, R. E. Rothman, D. Sullivan, S. Shoham, A. Casadevall, T. C. Quinn, A. Pekosz, A. A. R. Tobian, and O. Laeyendecker. 2020. Comparative performance of five commercially available serologic assays to detect antibodies to SARS-CoV-2 and identify individuals with high neutralizing titers. J Clin Microbiol.

21. Pisanic, N., P. R. Randad, K. Kruczynski, Y. C. Manabe, D. L. Thomas, A. Pekosz, S. L. Klein, M. J. Betenbaugh, W. A. Clarke, O. Laeyendecker, P. P. Caturegli, H. B. Larman, B. Detrick, J. K. Fairley, A. C. Sherman, N. Rouphael, S. Edupuganti, D. A. Granger, S. W. Granger, M. H. Collins, and C. D. Heaney. 2020. COVID-19 Serology at Population Scale: SARS-CoV-2-Specific Antibody Responses in Saliva. J Clin Microbiol 59.

22. Roda, A., S. Cavalera, F. Di Nardo, D. Calabria, S. Rosati, P. Simoni, B. Colitti, C. Baggiani, M. Roda, and L. Anfossi. 2021. Dual lateral flow optical/chemiluminescence immunosensors for the rapid detection of salivary and serum IgA in patients with COVID-19 disease. Biosens Bioelectron 172:112765.

23. Self, W. H., M. W. Tenforde, W. B. Stubblefield, L. R. Feldstein, J. S. Steingrub, N. Shapiro, A. A. Ginde, M. E. Prekker, S. M. Brown, I. D. Peltan, M. N. Gong, M. S. Aboodi, A. Khan, M. C. Exline, D. C. Files, K. W. Gibbs, C. J. Lindsell, T. W. Rice, I. D. Jones, N. Halasa, H. K. Talbot, C. G. Grijalva, J. D. Casey, D. N. Hager, N. Qadir, D. J. Henning, M. M. Coughlin, J. Schiffer, V. Semenova, H. Li, N. J. Thornburg, and M. M. Patel. 2020. Decline in SARS-CoV-2 Antibodies After Mild Infection Among Frontline Health Care Personnel in a Multistate Hospital Network - 12 States, April-August 2020. MMWR Morb Mortal Wkly Rep 69:1762–1766.

24. Silva, J., C. Lucas, M. Sundaram, B. Israelow, P. Wong, J. Klein, M. Tokuyama, P. Lu, A. Venkataraman, F. Liu, T. Mao, J. E. Oh, A. Park, A. Casanovas-Massana, C. B. F. Vogels, C. M. Muenker, J. Zell, J. B. Fournier, M. Campbell, M. Chiorazzi, E. Ruiz Fuentes, M. Petrone, C. C. Kalinich, I. M. Ott, A. Watkins, A. J. Moore, M. I. Nakahata, N. D. Grubaugh, S. Farhadian, C. Dela Cruz, A. Ko, W. L. Schulz, A. M. Ring, S. Ma, S. Omer, A. L. Wyllie, and A. Iwasaki. 2021. Saliva viral load is a dynamic unifying correlate of COVID-19 severity and mortality. medRxiv.

25. Sullivan, P. S., C. Sailey, J. L. Guest, J. Guarner, C. Kelley, A. J. Siegler, M. Valentine-Graves, L. Gravens, C. del Rio, and T. H. Sanchez. 2020. Detection of SARS-CoV-2 RNA and Antibodies in Diverse Samples: Protocol to Validate the Sufficiency of Provider-Observed, Home-Collected Blood, Saliva, and Oropharyngeal Samples. JMIR Public Health Surveill 6:e19054–e19054.

26. Tobian, A. A. R., and B. H. Shaz. 2020. Earlier the better: convalescent plasma. Blood 136:652–654.

27. Varadhachary, A., D. Chatterjee, J. Garza, R. P. Garr, C. Foley, A. F. Letkeman, J. Dean, D. Haug, J. Breeze, R. Traylor, A. Malek, R. Nath, and L. Linbeck. 2020. Salivary anti-SARS-CoV-2 IgA as an accessible biomarker of mucosal immunity against COVID-19. medRxiv.

28. Watkins, A. E., E. P. Fenichel, D. M. Weinberger, C. B. F. Vogels, D. E. Brackney, A. Casanovas-Massana, M. Campbell, J. Fournier, S. Bermejo, R. Datta, C. S. Dela Cruz, S. F. Farhadian, A. Iwasaki, A. I. Ko, N. D. Grubaugh, and A. L. Wyllie. 2020. Pooling saliva to increase SARS-CoV-2 testing capacity. medRxiv.

29. Wyllie, A. L., J. Fournier, A. Casanovas-Massana, M. Campbell, M. Tokuyama, P. Vijayakumar, J. L. Warren, B. Geng, M. C. Muenker, A. J. Moore, C. B. F. Vogels, M. E. Petrone, I. M. Ott, P. Lu, A. Venkataraman, A. Lu-Culligan, J. Klein, R. Earnest, M. Simonov, R. Datta, R. Handoko, N. Naushad, L. R. Sewanan, J. Valdez, E. B. White, S. Lapidus, C. C. Kalinich, X. Jiang, D. J. Kim, E. Kudo, M. Linehan, T. Mao, M. Moriyama, J. E. Oh, A. Park, J. Silva, E. Song, T. Takahashi, M. Taura, O. E. Weizman, P. Wong, Y. Yang, S. Bermejo, C. D. Odio, S. B. Omer, C. S. Dela Cruz, S. Farhadian, R. A. Martinello, A. Iwasaki, N. D. Grubaugh, and A. I. Ko. 2020. Saliva or Nasopharyngeal Swab Specimens for Detection of SARS-CoV-2. N Engl J Med 383:1283–1286.

