## Supplemental Tables 1 through 6 for "Comparative performance of multiplex salivary and commercially available serologic assays to detect SARS-CoV-2 IgG and neutralization titers"

**Supplemental Table 1. Description of SARS-CoV-2 antigens coupled to Luminex MagPlex beads.**

| **Source** | **Antigen** | **Cat. No.** |
| --- | --- | --- |
| GenScript | Nucleoprotein | Z03480 |
| The Native Antigen Company | Nucleoprotein | REC31851 |
| GenScript | Nucleoprotein | Z03480 |
| Sino Biological | Spike S1 RBD | 40592-V08H |
| GenScript | RBD (h) | Z03483 |
| Mt. Sinai | RBD | n/a |
| Mt. Sinai | Whole spike | n/a |
| Sino Biological | S1+S2 ECD | 40589-V08B1 |

**Supplemental Table 2. Characteristics of the SARS-CoV-2 convalescent individuals who provided both plasma and saliva samples included in the analytical sample.**

|  | **Overall, n=101^a^** |
| --- | --- |
| Median age (IQR) | 44 (34-56) |
| Male, n (%) | 43 (42.6%) |
| Race, n (%) |  |
| Non-Hispanic White | 73 (72.3%) |
| Non-Hispanic Black | 9 (8.9%) |
| Hispanic | 3 (3.0%) |
| Non-Hispanic Asian | 9 (8.9%) |
| Other | 7 (6.9%) |
| Hospitalized, n (%) | 15 (14.9%) |
| Median days since PCR+ test (IQR) | 50 (40-70) |

^a^Sample size that contributed to the complete case analysis.

**Supplemental Table 3. Concordance between each component of the multiplex SARS-Cov-2 antibody assay and the final algorithmic result of the multiplex salivary assay in COVID-19 convalescent individuals.**

| **Salivary assays^a,b^** | **Sum N/RBD/S S/CO, n = 101 (93+, 8-)** | | | |
| --- | --- | --- | --- | --- |
|  | **PPA** | **NPA** | **PA** | **Kappa** |
| GenScript N | 54.8% | 100.0% | 58.4% | 0.161 |
| NAC N | 89.2% | 100.0% | 90.1% | 0.568 |
| Sino Bio RBD | 100.0% | 87.5% | 99.0% | 0.928 |
| Mt. Sinai RBD | 90.3% | 100.0% | 91.1% | 0.597 |
| GenScript RBD | 95.7% | 100.0% | 96.0% | 0.779 |
| Sino Bio ECD | 96.8% | 87.5% | 96.0% | 0.756 |
| Mt. Sinai S | 49.5% | 100.0% | 53.5% | 0.134 |
| Sum N S/CO^c^ | 89.2% | 100.0% | 90.1% | 0.568 |
| Sum RBD S/CO^d^ | 96.8% | 100.0% | 97.0% | 0.826 |
| Sum S S/CO^e^ | 89.2% | 87.5% | 89.1% | 0.507 |

Note: PPA: Positive Percent Agreement; NPA: Negative Percent Agreement; PA: Percent Agreement; Kappa: Cohen's Kappa Coefficient; nAb: Neutralizing antibody; NAC: Native Antigen Company; Sino Bio: Sino Biological.

^a^Insufficient total IgG, instrument error, ID discrepancy or other exclusion reasons for salivary assay were set to missing.

^b^Each saliva assay result was considered positive, if median fluorescence intensity (MFI) above threshold, and considered negative if MFI below threshold.

^c^Sum N was considered positive if the sum of signal to cutoff ratios of 2 N antigens above threshold (mean plus 3 SD of pre-COVID samples).

^d^Sum RBD was considered positive if the sum of signal to cutoff ratios of 3 RBD antigens above threshold (mean plus 3 SD of pre-COVID samples).

^e^Sum S was considered positive if the sum of signal to cutoff ratios of 2 antigens above threshold (mean plus 3 SD of pre-COVID samples).

**Supplemental Table 4. Concordance between SARS-CoV-2 antibody tests of plasma from COVID-19 convalescent individuals.**

|  | **PPA** | **NPA** | **PA** | **Kappa** |
| --- | --- | --- | --- | --- |
|  | Ortho Vitros (88+, 13-) | | | |
| Euroimmun | 100.0% | 84.6% | 98.0% | 0.906 |
| BioRad | 95.5% | 61.5% | 91.1% | 0.589 |
| nAb | 75.0% | 76.9% | 75.2% | 0.320 |
|  | nAb (69+, 32-) | | | |
| Euroimmun | 98.6% | 31.3% | 77.2% | 0.362 |
| BioRad | 95.7% | 28.1% | 74.3% | 0.286 |
|  | Euroimmun (90+, 11-) | | | |
| BioRad | 94.4% | 63.6% | 91.1% | 0.559 |

Note: PPA: Positive Percent Agreement; NPA: Negative Percent Agreement; PA: Percent Agreement; Kappa: Cohen's Kappa Coefficient; nAb: Neutralizing antibody. nAb area under curve (AUC) <20 was considered as negative result, ≥20 as positive result. Indeterminate results for blood antibody tests were considered negative.

**Supplemental Table 5. Spearman’s rank correlation between the net median fluorescence intensity of multiplex salivary SARS-CoV-2 antibody assay and plasma serologic assay signal to cut-off ratio (S/CO) in COVID-19 convalescent individuals.**

| **Salivary assays^a^** | **Ortho Vitros S/CO** | | **Euroimmun AU** | | **BioRad OD** | | **nAb AUC** | |
| --- | --- | --- | --- | --- | --- | --- | --- | --- |
|  | **Spearman coefficient**  **(95% CI)^b^** | ***p* value** | **Spearman coefficient**  **(95% CI)^b^** | ***p* value** | **Spearman coefficient**  **(95% CI)^b^** | ***p* value** | **Spearman coefficient**  **(95% CI)^b^** | ***p* value** |
| GenScript N | 0.58 (0.42, 0.72) | <0.001 | 0.58 (0.43, 0.71) | <0.001 | 0.52 (0.34, 0.66) | <0.001 | 0.57 (0.41, 0.69) | <0.001 |
| NAC N | 0.63 (0.48, 0.75) | <0.001 | 0.62 (0.47, 0.74) | <0.001 | 0.57 (0.41, 0.71) | <0.001 | 0.63 (0.48, 0.74) | <0.001 |
| Sino Bio RBD | 0.85 (0.76, 0.91) | <0.001 | 0.83 (0.73, 0.89) | <0.001 | 0.41 (0.22, 0.57) | <0.001 | 0.77 (0.67, 0.84) | <0.001 |
| Mt. Sinai RBD | 0.83 (0.73, 0.90) | <0.001 | 0.80 (0.71, 0.87) | <0.001 | 0.39 (0.19, 0.55) | <0.001 | 0.75 (0.65, 0.83) | <0.001 |
| GenScript RBD | 0.86 (0.77, 0.92) | <0.001 | 0.83 (0.75, 0.89) | <0.001 | 0.40 (0.20, 0.56) | <0.001 | 0.77 (0.67, 0.85) | <0.001 |
| Sino Bio ECD | 0.63 (0.47, 0.75) | <0.001 | 0.61 (0.46, 0.73) | <0.001 | 0.38 (0.19, 0.54) | <0.001 | 0.62 (0.48, 0.73) | <0.001 |
| Mt. Sinai S | 0.81 (0.71, 0.88) | <0.001 | 0.79 (0.69, 0.86) | <0.001 | 0.44 (0.24, 0.59) | <0.001 | 0.76 (0.65, 0.84) | <0.001 |
| Sum N S/CO^c^ | 0.62 (0.46, 0.74) | <0.001 | 0.62 (0.46, 0.73) | <0.001 | 0.57 (0.41, 0.71) | <0.001 | 0.62 (0.47, 0.72) | <0.001 |
| Sum RBD S/CO^d^ | 0.86 (0.76, 0.91) | <0.001 | 0.83 (0.74, 0.89) | <0.001 | 0.41 (0.22, 0.57) | <0.001 | 0.77 (0.66, 0.85) | <0.001 |
| Sum S S/CO^e^ | 0.68 (0.53, 0.79) | <0.001 | 0.66 (0.52, 0.77) | <0.001 | 0.40 (0.20, 0.56) | <0.001 | 0.67 (0.54, 0.77) | <0.001 |
| Sum N/S/RBD S/CO^f^ | 0.80 (0.69, 0.87) | <0.001 | 0.77 (0.68, 0.85) | <0.001 | 0.44 (0.26, 0.59) | <0.001 | 0.75 (0.65, 0.83) | <0.001 |

Note: S/CO = signal to cut off ratio. AU = arbitrary units. OD = optical density. nAb = neutralizing antibody. AUC: area under curve. CI: confidence interval. NAC: Native Antigen Company. Sino Bio: Sino Biological.

^a^Salivary assay values were signal to cutoff.

^b^Spearman’s correlation coefficients (ρ) were calculated with 95% CI estimated over 1000 bootstrap iterations.

^c^Sum of S/CO of 2 N antigens.

^d^Sum of S/CO of 3 RBD antigens.

^e^Sum of S/CO of 2 Spike antigens.

^f^Sum of 7 N, RBD and S antigen S/CO.

**Supplemental Table 6. Spearman’s rank correlation between SARS-CoV-2 antibody tests of plasma from COVID-19 convalescent individuals.**

|  | **Spearman coefficient (95% CI)** | ***p* value** |
| --- | --- | --- |
|  | Ortho, S/CO | |
| Euroimmun AU | 0.93 (0.90, 0.95) | <0.001 |
| BioRad OD | 0.43 (0.23, 0.59) | <0.001 |
| nAb AUC | 0.83 (0.74, 0.89) | <0.001 |
|  | nAb | |
| Euroimmun AU | 0.80 (0.72, 0.86) | <0.001 |
| BioRad OD | 0.38 (0.18, 0.55) | <0.001 |
|  | Euroimmun AU | |
| BioRad OD | 0.45 (0.26, 0.61) | <0.001 |

Note: S/CO = signal to cut off ratio. AU = arbitrary units. OD = optical density. nAb = neutralizing antibody. AUC: area under curve. CI: confidence interval.

Spearman’s correlation coefficients (ρ) were calculated with 95% CI estimated over 1000 bootstrap iterations.
